# Temporal trends in hospitalizations and 30-day mortality in older patients during the COVID pandemic from March 2020 to July 2021

**DOI:** 10.1101/2021.12.22.21268237

**Authors:** Sara Garcia-Ptacek, Hong Xu, Martin Annetorp, Viktoria Bäck Jerlardtz, Tommy Cederholm, Malin Engström, Miia Kivipelto, Lars Göran Lundberg, Carina Metzner, Maria Olsson, Josefina Skogö Nyvang, Carina Sühl Öberg, Elisabet Åkesson, Dorota Religa, Maria Eriksdotter

## Abstract

**Importance:** Previous reports have suggested reductions in mortality risk from COVID-19 throughout the first wave of the COVID-19 pandemic. Mortality changes later in the pandemic and pandemic effects on other types of geriatric hospitalizations are less studied.

**Objectives:** To describe the changes in hospitalizations and 30-day mortality in Stockholm for patients 70+ receiving inpatient geriatric care for COVID-19 and other causes.

**Design:** Observational study. For patients 70 or older, we present the incidence of 30-day mortality from COVID-19 in the Stockholm region, in relationship to geriatric hospitalizations and 30-day mortality after admission for COVID-19 and other causes.

**Setting:** Hospitalizations for patients 70+ from geriatric clinics in Stockholm, Sweden hospitalized for COVID-19 or other causes between March 2020 and July 31, 2021, were included.

**Participants:** The total number of geriatric hospitalizations for patients 70+ was 5,320 for COVID-19 and 32,243 for non-COVID-19 causes, corresponding to 4,565 individual COVID-19 patients and 19,308 non-COVID-19 patients.

**Exposure(s):** The date of hospital admission to a geriatric clinic. Main Outcome(s) and Measure(s): 30-day mortality after admission.

**Results:** In patients with COVID-19, the 30-day mortality rate was highest at the beginning of the first wave (29% in March-April 2020), decreased as the first wave subsided (7% July-August), increased again in the second wave (17% November-December), but failed to increase as much in the third wave (11-13% March-July 2021). In non-COVID-19 geriatric patients during the same period, the 30-day mortality presented a similar trend, but with a smaller magnitude of variation (5 to 10%). The number of persons 70 or older testing positive for COVID-19 in Stockholm reached two peaks in 2020 (April and December), fell in January 2021 and then increased again in March-April 2021.

**Conclusions and Relevance:** During the first and second waves, hospital admissions and 30-day mortality after geriatric hospitalization for COVID-19 increased in periods of high community transmission, although the mortality peak was lower in wave 2 than in wave 1. The mortality for non-COVID geriatric cases was lower and more stable but also showed an increase with the pandemic peaks.

**KEY POINTS:** *Question:* Multiple previous reports in different countries and settings have shown higher case fatality ratio or hospitalized case fatality ratio for COVID-19 in the first wave compared to the second wave of the pandemic. However, less is known about how the COVID-19 waves specifically affected the care of geriatric patients, including those with conditions other than COVID.

*Findings:* The total number of hospitalizations was 5,320 for COVID-19 and 32,243 for non-COVID-cases. In COVID-patients, the 30-day mortality rate was highest at the beginning of the first wave (29% in March-April 2020), reached 17% at the second wave peak (November-December) followed by 11-13% in the third wave (March-July 2021). The mortality in non-COVID geriatric patients showed a similar trend, but of lower magnitude (5-10%). During the incidence peaks, COVID-19 hospitalizations displaced non-COVID geriatric patients.

*Meaning:* Hospital admissions and 30-day mortality after hospitalizations for COVID-19 increased in periods of high community transmission, albeit with decreasing mortality rates from wave 1 to 3, with a possible vaccination effect in wave 3. Thus, the healthcare system could not compensate for the high community spread of COVID-19 during the pandemic peaks, which also led to displacing care for non-COVID geriatric patients. These results are important for planning healthcare resources in future health emergencies.

## INTRODUCTION

During the first wave of the COVID-19 pandemic in Stockholm from March to July 2020, mortality for COVID-19 patients hospitalized in geriatric clinics decreased over time.^1^ It was unclear at the time whether this was due to improved care or to patient selection of more severe cases earlier in the pandemic due to higher community incidence and an overburdened healthcare system. Our results from that first wave included only in-hospital mortality and did not explore how care for conditions other than COVID-19 was impacted by the pandemic. Age is among the strongest risk factors for COVID-19 and the infection fatality ratio after age 70 is substantial.^2, 3^ Here, we present data on geriatric hospitalizations for COVID-19 and other causes, including 30-day mortality from admission and compare these to the incidence of COVID-19 and 30-day mortality from COVID-19 in the Stockholm region for persons 70+. We aim to describe how COVID-19 peaks affected geriatric patients 70 and older hospitalized both for COVID-19 and for other causes in Stockholm.^4-7^

In previous reports, COVID-19 mortality in hospitalized patients in Sweden increased with each pandemic wave and decreased between pandemic peaks.^8, 9^ According to the Swedish Board of Health and Welfare, 60-day mortality after admission with COVID-19 decreased from a first-wave peak in March 2020 (25%) to a first nadir in August-September 2020 (10%), then rose again but to lower levels as the second wave progressed to a peak in December 2020 (20%), decreasing steadily afterwards.^10^ Decreasing mortality during the first pandemic wave was also observed in studies from the US,^11^ with subsequent increases in mortality during pandemic surges.^12^ A Spanish study reported that COVID-19 patients in the first wave were older, with more comorbidities and higher mortality than in the second wave.^13^ However, information is missing on mortality trends later in the pandemic, trends in older population and in geriatric patients with diagnoses other than COVID-19.

The aim of this study is to describe the changes over time in 30 day-mortality after admission of patients 70 years and older, hospitalized in 9 geriatric clinics in the Stockholm region from March 2020 to July 2021. We describe 30-day mortality both for patients hospitalized for COVID-19 and other causes over the three waves of the pandemic. Weekly hospitalizations for COVID-19 and other causes are presented and compared to the average weekly hospitalizations in the same geriatric clinics in 2019. These results are presented in the context of COVID-19 incidence and 30-day mortality and weekly hospitalizations from any cause in population 70 and over in the Stockholm region.

## METHODS

### Study population

We identified all hospitalizations of patients 70 years and older who were admitted to nine geriatric hospitals in Stockholm, Sweden, from March 6^th^, 2020, to July 31st, 2021. We excluded hospitalizations with a duration less than 24 hours or with an admission date after August 1^st^, 2021, to allow one month follow-up for mortality. A total of 5,320 hospitalizations for COVID-19 were included, together with 32,243 hospitalizations with non-COVID-19 diagnoses during the same period (**Figure 1**). This corresponded to 4,565 individual COVID-19 patients and 19,308 non-COVID-19 patients.

**Figure 1.**
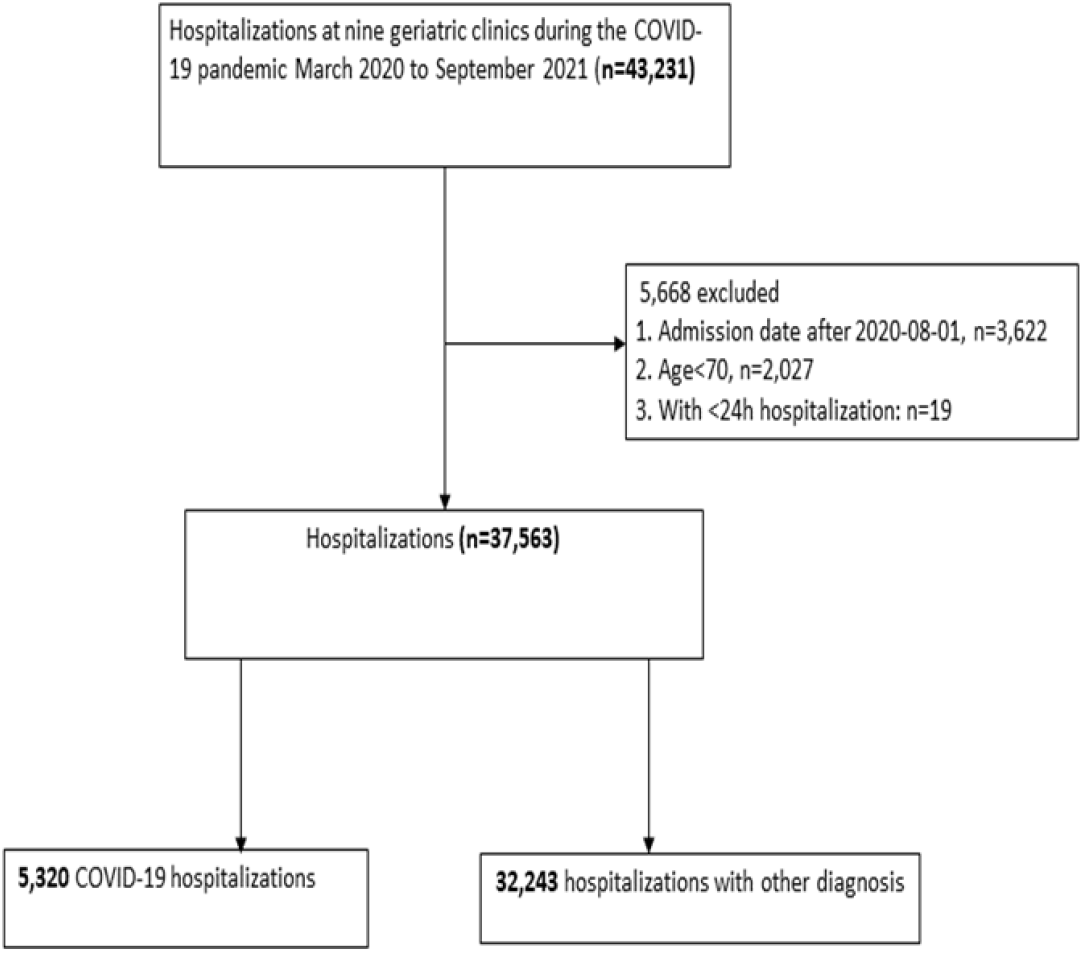
Case selection flow chart.

### Exposure and Outcome

The study exposure was the hospital admission date (March 1^st^, 2020, to July 31^st^, 2021). The study outcome was 30-day mortality from admission. Patients were censored at death, or the end of follow-up (September 10th, 2021) whichever came first.

### COVID-19 diagnosis and covariates

The diagnosis of COVID-19 followed clinical practice and was based on a positive reverse transcriptase-polymerase chain reaction (RT-PCR) analysis from nasopharyngeal swabs or, in case of a negative RT-PCR, typical clinical picture (including a consultation with a specialist in infectious diseases) and a CT scan with typical COVID-19 findings. All hospitalizations were included, and patients could have multiple hospitalizations. Hospitalizations for diagnoses other than COVID-19 to the same nine geriatric hospitals in Stockholm during the same period comprised the non-COVID group. We collected information on patient demographics, initial vital signs, medications, diagnoses at discharge, and 30-day mortality through the hospital electronic health records. 30-day mortality was defined from the date of admission to geriatrics.

### COVID-19 incidence and mortality in the population 70 years and older in Stockholm

Information on confirmed cases with COVID-19 and mortality data in the population 70 and over in Stockholm was provided by the Swedish Board of Health and Welfare for research purposes (www.socialstyrelsen.se). Incidence data represents detected COVID-19 infections. Thirty-day mortality corresponds to deaths where COVID-19 was the main cause of death within 30 days of a positive test for persons 70 and over in the Stockholm region. Weekly hospitalizations for any cause in this population were also included as reference.

### Weekly hospitalizations in 2019

For comparison, the average weekly hospitalizations registered in 2019 in the 9 geriatric clinics was presented. This was calculated by adding the total number of hospitalizations from all clinics (30,969) and dividing by 52 calendar weeks resulting in an average of 596 new hospitalizations per week.

### Analysis

Summary statistics are displayed as mean ± standard deviation (SD) or median (interquartile range, IQR) or proportions.

30-day mortality rate is graphically presented in relationship to number of new admissions per week and COVID-19 incidence and mortality in the Stockholm region. Via logistic regression models, we assessed the effect of admission date on 30-day mortality, adjusted by age, sex, and medical treatment. Results were reported as odds ratios (OR) and 95% confidence interval (CI). Mortality rates were also compared against non-COVID-19 patients admitted to the same geriatric clinics during the same period.

All analyses were performed using R (https://www.r-project.org) and Stata version 17.0 (StataCorp, College Station, TX).

### Ethical statement

The Swedish Ethical Review Authority approved the study (Dnr 2020-02146, and 2020-03345).

## RESULTS

### Characteristics and treatments at admission

Among the hospitalizations due to COVID-19, 87 % (4645 hospitalizations) had positive COVID-19 RT-PCR while 675 (13%) had a negative RT-PCR but fulfilled the clinical and radiological criteria for COVID-19. Patients hospitalized for COVID-19 had a median age of 84 (IQR 78-89) years, 53% were women and 6% of the patients had low saturation (<90%) at admission. The median duration of hospitalization was 9 days (IQR 6-13) (**Table 1**). The median age of geriatric hospitalizations for other (non-COVID-19) diagnoses was higher (85 IQR 79-90), and their hospital stays were shorter (6 days: IQR 4-9).

**Table 1.**
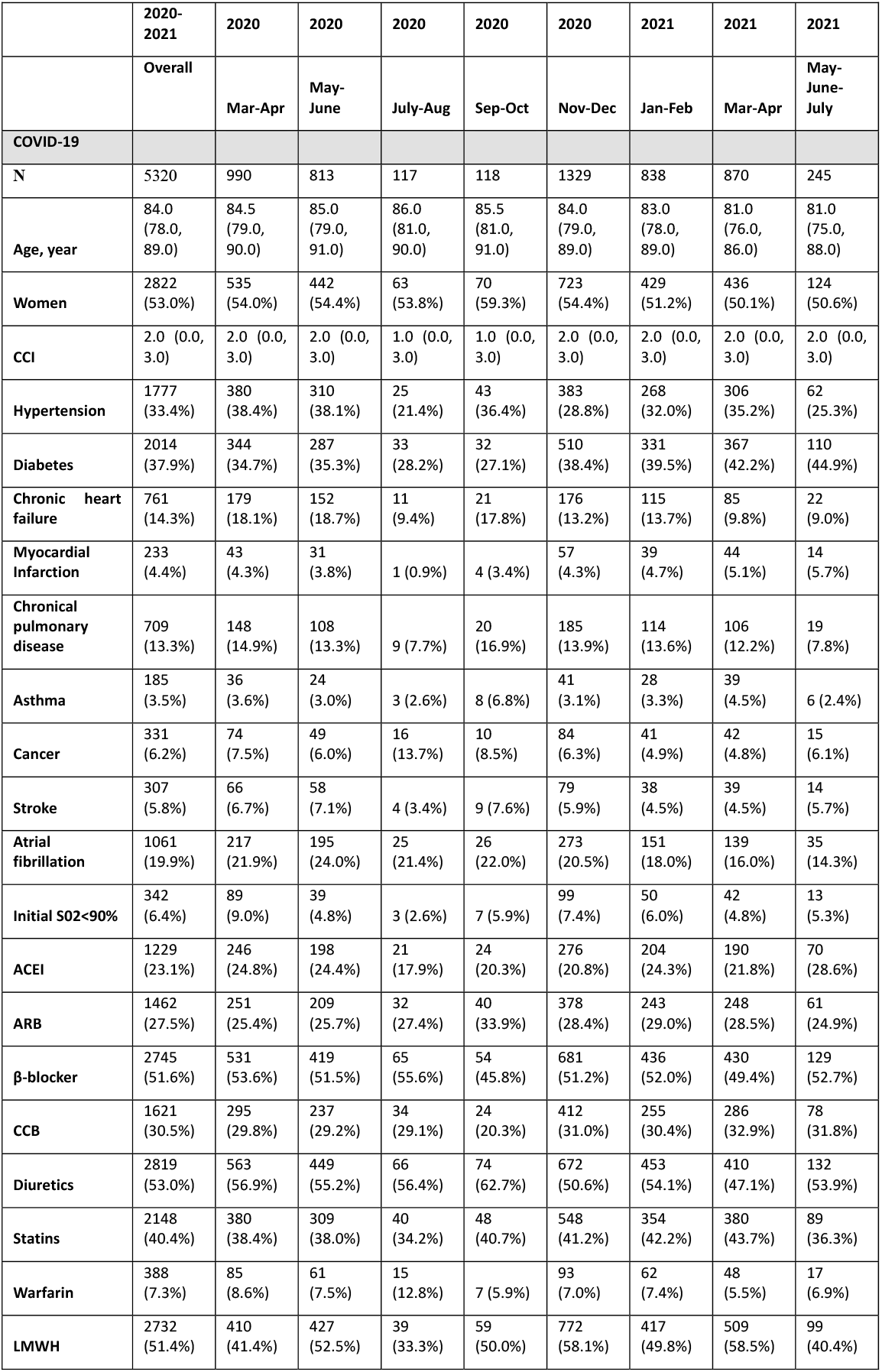
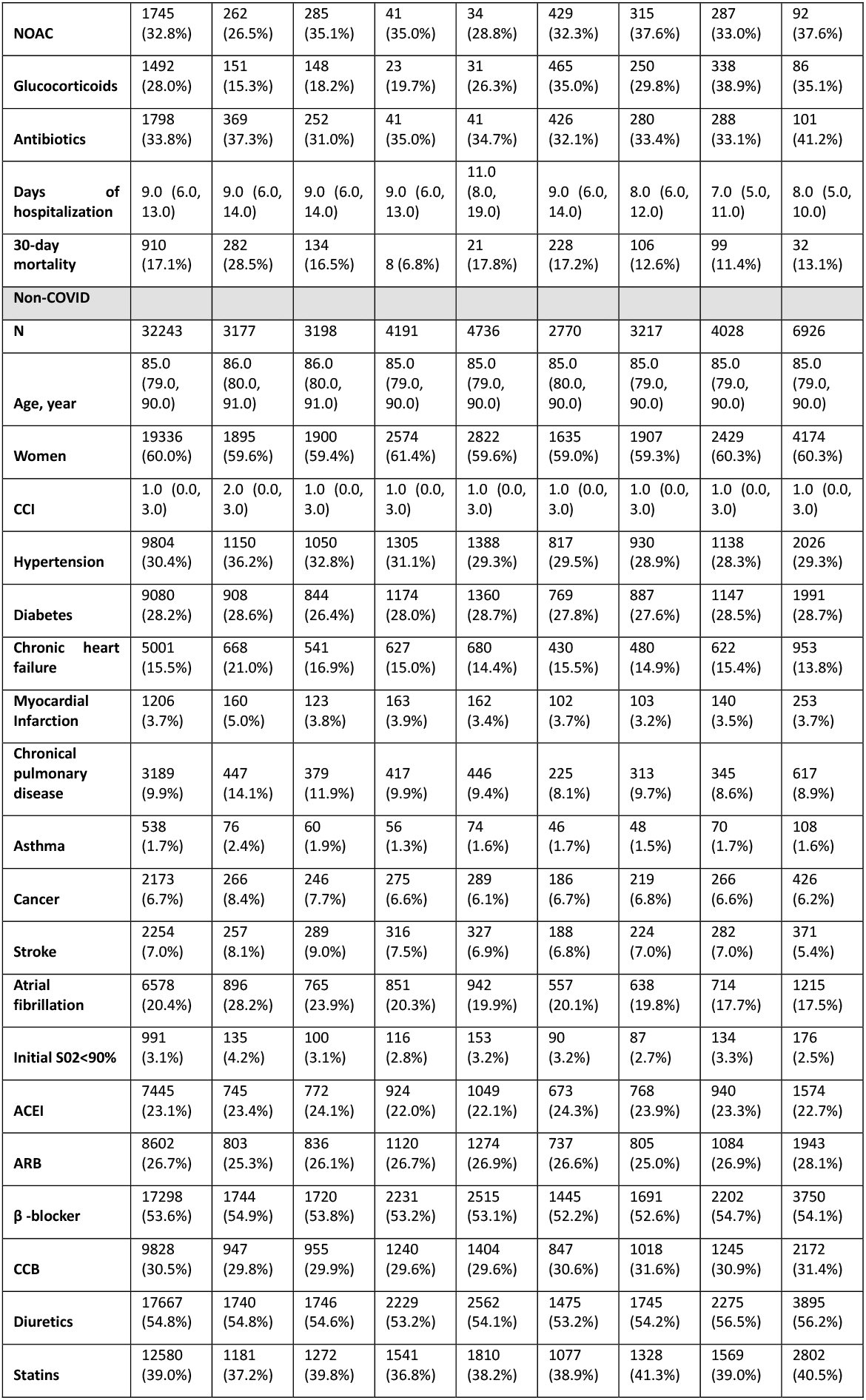
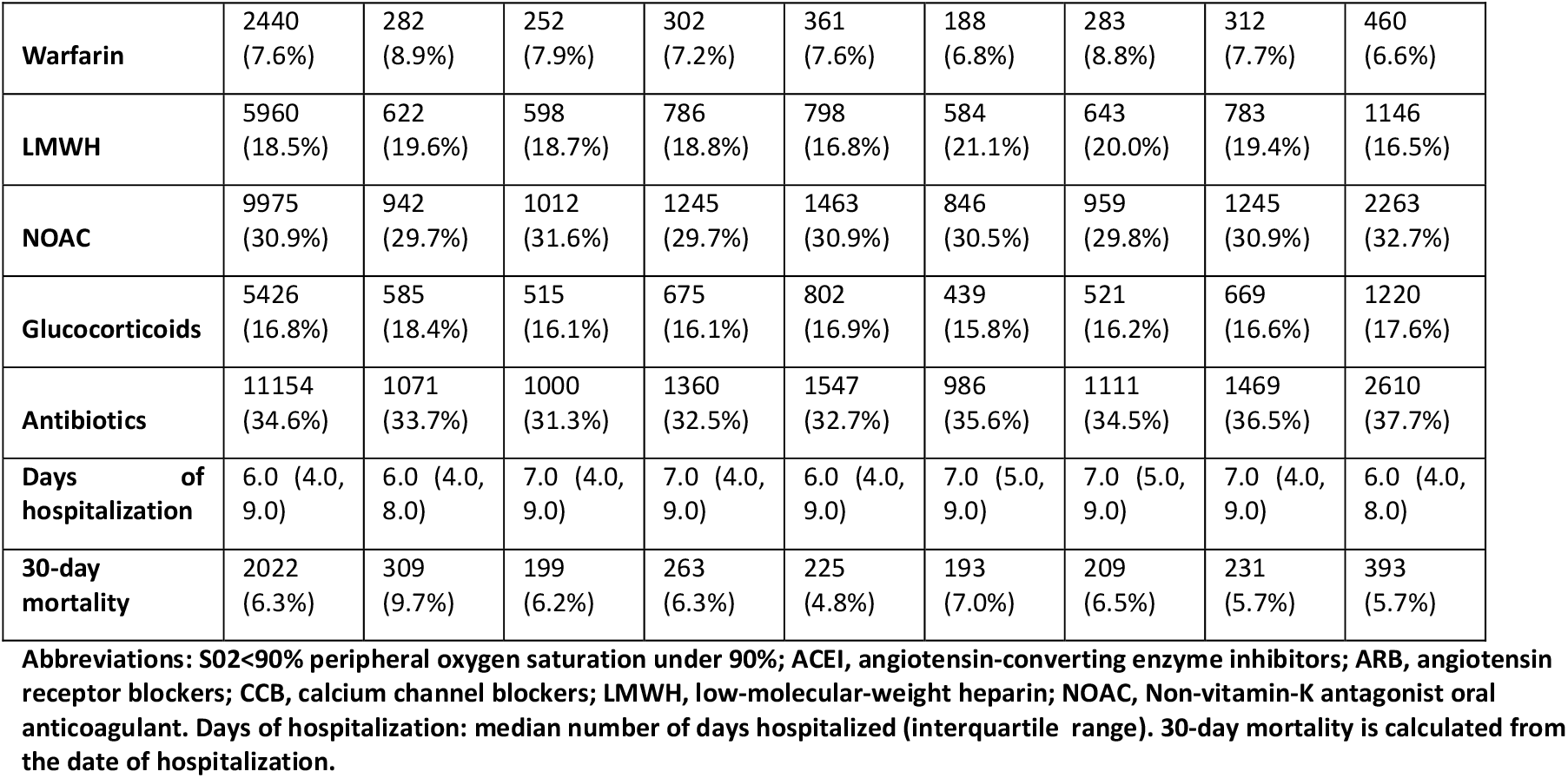
Hospitalizations for COVID-19 and other causes in patients 70 and over in geriatric clinics.

|  | 2020-2021 | 2020 | 2020 | 2020 | 2020 | 2020 | 2021 | 2021 | 2021 |
| --- | --- | --- | --- | --- | --- | --- | --- | --- | --- |
|  | Overall | Mar-Apr | May-June | July-Aug | Sep-Oct | Nov-Dec | Jan-Feb | Mar-Apr | May-June-July |
| **COVID-19** |  |  |  |  |  |  |  |  |  |
| N | 5320 | 990 | 813 | 117 | 118 | 1329 | 838 | 870 | 245 |
| Age, year | 84.0<br>(78.0, 89.0) | 84.5<br>(79.0, 90.0) | 85.0<br>(79.0, 91.0) | 86.0<br>(81.0, 90.0) | 85.5<br>(81.0, 91.0) | 84.0<br>(79.0, 89.0) | 83.0<br>(78.0, 89.0) | 81.0<br>(76.0, 86.0) | 81.0<br>(75.0, 88.0) |
| Women | 2822<br>(53.0%) | 535<br>(54.0%) | 442<br>(54.4%) | 63<br>(53.8%) | 70<br>(59.3%) | 723<br>(54.4%) | 429<br>(51.2%) | 436<br>(50.1%) | 124<br>(50.6%) |
| CCI | 2.0 (0.0, 3.0) | 2.0 (0.0, 3.0) | 2.0 (0.0, 3.0) | 1.0 (0.0, 3.0) | 1.0 (0.0, 3.0) | 2.0 (0.0, 3.0) | 2.0 (0.0, 3.0) | 2.0 (0.0, 3.0) | 2.0 (0.0, 3.0) |
| Hypertension | 1777<br>(33.4%) | 380<br>(38.4%) | 310<br>(38.1%) | 25<br>(21.4%) | 43<br>(36.4%) | 383<br>(28.8%) | 268<br>(32.0%) | 306<br>(35.2%) | 62<br>(25.3%) |
| Diabetes | 2014<br>(37.9%) | 344<br>(34.7%) | 287<br>(35.3%) | 33<br>(28.2%) | 32<br>(27.1%) | 510<br>(38.4%) | 331<br>(39.5%) | 367<br>(42.2%) | 110<br>(44.9%) |
| Chronic heart failure | 761<br>(14.3%) | 179<br>(18.1%) | 152<br>(18.7%) | 11<br>(9.4%) | 21<br>(17.8%) | 176<br>(13.2%) | 115<br>(13.7%) | 85<br>(9.8%) | 22<br>(9.0%) |
| Myocardial Infarction | 233<br>(4.4%) | 43<br>(4.3%) | 31<br>(3.8%) | 1 (0.9%) | 4 (3.4%) | 57<br>(4.3%) | 39<br>(4.7%) | 44<br>(5.1%) | 14<br>(5.7%) |
| Chronical pulmonary disease | 709<br>(13.3%) | 148<br>(14.9%) | 108<br>(13.3%) | 9 (7.7%) | 20<br>(16.9%) | 185<br>(13.9%) | 114<br>(13.6%) | 106<br>(12.2%) | 19<br>(7.8%) |
| Asthma | 185<br>(3.5%) | 36<br>(3.6%) | 24<br>(3.0%) | 3 (2.6%) | 8 (6.8%) | 41<br>(3.1%) | 28<br>(3.3%) | 39<br>(4.5%) | 6 (2.4%) |
| Cancer | 331<br>(6.2%) | 74<br>(7.5%) | 49<br>(6.0%) | 16<br>(13.7%) | 10<br>(8.5%) | 84<br>(6.3%) | 41<br>(4.9%) | 42<br>(4.8%) | 15<br>(6.1%) |
| Stroke | 307<br>(5.8%) | 66<br>(6.7%) | 58<br>(7.1%) | 4 (3.4%) | 9 (7.6%) | 79<br>(5.9%) | 38<br>(4.5%) | 39<br>(4.5%) | 14<br>(5.7%) |
| Atrial fibrillation | 1061<br>(19.9%) | 217<br>(21.9%) | 195<br>(24.0%) | 25<br>(21.4%) | 26<br>(22.0%) | 273<br>(20.5%) | 151<br>(18.0%) | 139<br>(16.0%) | 35<br>(14.3%) |
| Initial S02<90% | 342<br>(6.4%) | 89<br>(9.0%) | 39<br>(4.8%) | 3 (2.6%) | 7 (5.9%) | 99<br>(7.4%) | 50<br>(6.0%) | 42<br>(4.8%) | 13<br>(5.3%) |
| ACEI | 1229<br>(23.1%) | 246<br>(24.8%) | 198<br>(24.4%) | 21<br>(17.9%) | 24<br>(20.3%) | 276<br>(20.8%) | 204<br>(24.3%) | 190<br>(21.8%) | 70<br>(28.6%) |
| ARB | 1462<br>(27.5%) | 251<br>(25.4%) | 209<br>(25.7%) | 32<br>(27.4%) | 40<br>(33.9%) | 378<br>(28.4%) | 243<br>(29.0%) | 248<br>(28.5%) | 61<br>(24.9%) |
| β-blocker | 2745<br>(51.6%) | 531<br>(53.6%) | 419<br>(51.5%) | 65<br>(55.6%) | 54<br>(45.8%) | 681<br>(51.2%) | 436<br>(52.0%) | 430<br>(49.4%) | 129<br>(52.7%) |
| CCB | 1621<br>(30.5%) | 295<br>(29.8%) | 237<br>(29.2%) | 34<br>(29.1%) | 24<br>(20.3%) | 412<br>(31.0%) | 255<br>(30.4%) | 286<br>(32.9%) | 78<br>(31.8%) |
| Diuretics | 2819<br>(53.0%) | 563<br>(56.9%) | 449<br>(55.2%) | 66<br>(56.4%) | 74<br>(62.7%) | 672<br>(50.6%) | 453<br>(54.1%) | 410<br>(47.1%) | 132<br>(53.9%) |
| Statins | 2148<br>(40.4%) | 380<br>(38.4%) | 309<br>(38.0%) | 40<br>(34.2%) | 48<br>(40.7%) | 548<br>(41.2%) | 354<br>(42.2%) | 380<br>(43.7%) | 89<br>(36.3%) |
| Warfarin | 388<br>(7.3%) | 85<br>(8.6%) | 61<br>(7.5%) | 15<br>(12.8%) | 7 (5.9%) | 93<br>(7.0%) | 62<br>(7.4%) | 48<br>(5.5%) | 17<br>(6.9%) |
| LMWH | 2732<br>(51.4%) | 410<br>(41.4%) | 427<br>(52.5%) | 39<br>(33.3%) | 59<br>(50.0%) | 772<br>(58.1%) | 417<br>(49.8%) | 509<br>(58.5%) | 99<br>(40.4%) |
| **NOAC** | 1745<br>(32.8%) | 262<br>(26.5%) | 285<br>(35.1%) | 41<br>(35.0%) | 34<br>(28.8%) | 429<br>(32.3%) | 315<br>(37.6%) | 287<br>(33.0%) | 92<br>(37.6%) |
| **Glucocorticoids** | 1492<br>(28.0%) | 151<br>(15.3%) | 148<br>(18.2%) | 23<br>(19.7%) | 31<br>(26.3%) | 465<br>(35.0%) | 250<br>(29.8%) | 338<br>(38.9%) | 86<br>(35.1%) |
| **Antibiotics** | 1798<br>(33.8%) | 369<br>(37.3%) | 252<br>(31.0%) | 41<br>(35.0%) | 41<br>(34.7%) | 426<br>(32.1%) | 280<br>(33.4%) | 288<br>(33.1%) | 101<br>(41.2%) |
| **Days of hospitalization** | 9.0 (6.0, 13.0) | 9.0 (6.0, 14.0) | 9.0 (6.0, 14.0) | 9.0 (6.0, 13.0) | 11.0 (8.0, 19.0) | 9.0 (6.0, 14.0) | 8.0 (6.0, 12.0) | 7.0 (5.0, 11.0) | 8.0 (5.0, 10.0) |
| **30-day mortality** | 910<br>(17.1%) | 282<br>(28.5%) | 134<br>(16.5%) | 8 (6.8%) | 21<br>(17.8%) | 228<br>(17.2%) | 106<br>(12.6%) | 99<br>(11.4%) | 32<br>(13.1%) |
| **Non-COVID** |  |  |  |  |  |  |  |  |  |
| **N** | 32243 | 3177 | 3198 | 4191 | 4736 | 2770 | 3217 | 4028 | 6926 |
| **Age, year** | 85.0<br>(79.0, 90.0) | 86.0<br>(80.0, 91.0) | 86.0<br>(80.0, 91.0) | 85.0<br>(79.0, 90.0) | 85.0<br>(79.0, 90.0) | 85.0<br>(80.0, 90.0) | 85.0<br>(79.0, 90.0) | 85.0<br>(79.0, 90.0) | 85.0<br>(79.0, 90.0) |
| **Women** | 19336<br>(60.0%) | 1895<br>(59.6%) | 1900<br>(59.4%) | 2574<br>(61.4%) | 2822<br>(59.6%) | 1635<br>(59.0%) | 1907<br>(59.3%) | 2429<br>(60.3%) | 4174<br>(60.3%) |
| **CCI** | 1.0 (0.0, 3.0) | 2.0 (0.0, 3.0) | 1.0 (0.0, 3.0) | 1.0 (0.0, 3.0) | 1.0 (0.0, 3.0) | 1.0 (0.0, 3.0) | 1.0 (0.0, 3.0) | 1.0 (0.0, 3.0) | 1.0 (0.0, 3.0) |
| **Hypertension** | 9804<br>(30.4%) | 1150<br>(36.2%) | 1050<br>(32.8%) | 1305<br>(31.1%) | 1388<br>(29.3%) | 817<br>(29.5%) | 930<br>(28.9%) | 1138<br>(28.3%) | 2026<br>(29.3%) |
| **Diabetes** | 9080<br>(28.2%) | 908<br>(28.6%) | 844<br>(26.4%) | 1174<br>(28.0%) | 1360<br>(28.7%) | 769<br>(27.8%) | 887<br>(27.6%) | 1147<br>(28.5%) | 1991<br>(28.7%) |
| **Chronic heart failure** | 5001<br>(15.5%) | 668<br>(21.0%) | 541<br>(16.9%) | 627<br>(15.0%) | 680<br>(14.4%) | 430<br>(15.5%) | 480<br>(14.9%) | 622<br>(15.4%) | 953<br>(13.8%) |
| **Myocardial Infarction** | 1206<br>(3.7%) | 160<br>(5.0%) | 123<br>(3.8%) | 163<br>(3.9%) | 162<br>(3.4%) | 102<br>(3.7%) | 103<br>(3.2%) | 140<br>(3.5%) | 253<br>(3.7%) |
| **Chronical pulmonary disease** | 3189<br>(9.9%) | 447<br>(14.1%) | 379<br>(11.9%) | 417<br>(9.9%) | 446<br>(9.4%) | 225<br>(8.1%) | 313<br>(9.7%) | 345<br>(8.6%) | 617<br>(8.9%) |
| **Asthma** | 538<br>(1.7%) | 76<br>(2.4%) | 60<br>(1.9%) | 56<br>(1.3%) | 74<br>(1.6%) | 46<br>(1.7%) | 48<br>(1.5%) | 70<br>(1.7%) | 108<br>(1.6%) |
| **Cancer** | 2173<br>(6.7%) | 266<br>(8.4%) | 246<br>(7.7%) | 275<br>(6.6%) | 289<br>(6.1%) | 186<br>(6.7%) | 219<br>(6.8%) | 266<br>(6.6%) | 426<br>(6.2%) |
| **Stroke** | 2254<br>(7.0%) | 257<br>(8.1%) | 289<br>(9.0%) | 316<br>(7.5%) | 327<br>(6.9%) | 188<br>(6.8%) | 224<br>(7.0%) | 282<br>(7.0%) | 371<br>(5.4%) |
| **Atrial fibrillation** | 6578<br>(20.4%) | 896<br>(28.2%) | 765<br>(23.9%) | 851<br>(20.3%) | 942<br>(19.9%) | 557<br>(20.1%) | 638<br>(19.8%) | 714<br>(17.7%) | 1215<br>(17.5%) |
| **Initial S02<90%** | 991<br>(3.1%) | 135<br>(4.2%) | 100<br>(3.1%) | 116<br>(2.8%) | 153<br>(3.2%) | 90<br>(3.2%) | 87<br>(2.7%) | 134<br>(3.3%) | 176<br>(2.5%) |
| **ACEI** | 7445<br>(23.1%) | 745<br>(23.4%) | 772<br>(24.1%) | 924<br>(22.0%) | 1049<br>(22.1%) | 673<br>(24.3%) | 768<br>(23.9%) | 940<br>(23.3%) | 1574<br>(22.7%) |
| **ARB** | 8602<br>(26.7%) | 803<br>(25.3%) | 836<br>(26.1%) | 1120<br>(26.7%) | 1274<br>(26.9%) | 737<br>(26.6%) | 805<br>(25.0%) | 1084<br>(26.9%) | 1943<br>(28.1%) |
| **β -blocker** | 17298<br>(53.6%) | 1744<br>(54.9%) | 1720<br>(53.8%) | 2231<br>(53.2%) | 2515<br>(53.1%) | 1445<br>(52.2%) | 1691<br>(52.6%) | 2202<br>(54.7%) | 3750<br>(54.1%) |
| **CCB** | 9828<br>(30.5%) | 947<br>(29.8%) | 955<br>(29.9%) | 1240<br>(29.6%) | 1404<br>(29.6%) | 847<br>(30.6%) | 1018<br>(31.6%) | 1245<br>(30.9%) | 2172<br>(31.4%) |
| **Diuretics** | 17667<br>(54.8%) | 1740<br>(54.8%) | 1746<br>(54.6%) | 2229<br>(53.2%) | 2562<br>(54.1%) | 1475<br>(53.2%) | 1745<br>(54.2%) | 2275<br>(56.5%) | 3895<br>(56.2%) |
| **Statins** | 12580<br>(39.0%) | 1181<br>(37.2%) | 1272<br>(39.8%) | 1541<br>(36.8%) | 1810<br>(38.2%) | 1077<br>(38.9%) | 1328<br>(41.3%) | 1569<br>(39.0%) | 2802<br>(40.5%) |
| **Warfarin** | 2440<br>(7.6%) | 282<br>(8.9%) | 252<br>(7.9%) | 302<br>(7.2%) | 361<br>(7.6%) | 188<br>(6.8%) | 283<br>(8.8%) | 312<br>(7.7%) | 460<br>(6.6%) |
| **LMWH** | 5960<br>(18.5%) | 622<br>(19.6%) | 598<br>(18.7%) | 786<br>(18.8%) | 798<br>(16.8%) | 584<br>(21.1%) | 643<br>(20.0%) | 783<br>(19.4%) | 1146<br>(16.5%) |
| **NOAC** | 9975<br>(30.9%) | 942<br>(29.7%) | 1012<br>(31.6%) | 1245<br>(29.7%) | 1463<br>(30.9%) | 846<br>(30.5%) | 959<br>(29.8%) | 1245<br>(30.9%) | 2263<br>(32.7%) |
| **Glucocorticoids** | 5426<br>(16.8%) | 585<br>(18.4%) | 515<br>(16.1%) | 675<br>(16.1%) | 802<br>(16.9%) | 439<br>(15.8%) | 521<br>(16.2%) | 669<br>(16.6%) | 1220<br>(17.6%) |
| **Antibiotics** | 11154<br>(34.6%) | 1071<br>(33.7%) | 1000<br>(31.3%) | 1360<br>(32.5%) | 1547<br>(32.7%) | 986<br>(35.6%) | 1111<br>(34.5%) | 1469<br>(36.5%) | 2610<br>(37.7%) |
| **Days of hospitalization** | 6.0 (4.0, 9.0) | 6.0 (4.0, 8.0) | 7.0 (4.0, 9.0) | 7.0 (4.0, 9.0) | 6.0 (4.0, 9.0) | 7.0 (5.0, 9.0) | 7.0 (5.0, 9.0) | 7.0 (4.0, 9.0) | 6.0 (4.0, 8.0) |
| **30-day mortality** | 2022<br>(6.3%) | 309<br>(9.7%) | 199<br>(6.2%) | 263<br>(6.3%) | 225<br>(4.8%) | 193<br>(7.0%) | 209<br>(6.5%) | 231<br>(5.7%) | 393<br>(5.7%) |

Abbreviations: S02<90% peripheral oxygen saturation under 90%; ACEI, angiotensin-converting enzyme inhibitors; ARB, angiotensin receptor blockers; CCB, calcium channel blockers; LMWH, low-molecular-weight heparin; NOAC, Non-vitamin-K antagonist oral anticoagulant. Days of hospitalization: median number of days hospitalized (interquartile range). 30-day mortality is calculated from the date of hospitalization.

The total number of hospitalizations in the geriatric hospitals fell sharply at the beginning of the pandemic from a pre-pandemic weekly average of 596 in 2019. After this initial drop in the first wave, geriatric hospitalizations rallied, averaging 507 hospitalizations per week but without a sustained return to pre-pandemic levels. When COVID-19 cases increased, non-COVID-19 cases decreased accordingly (**Figure 2A**; **Supplemental table 1**).

**Figure 2.**
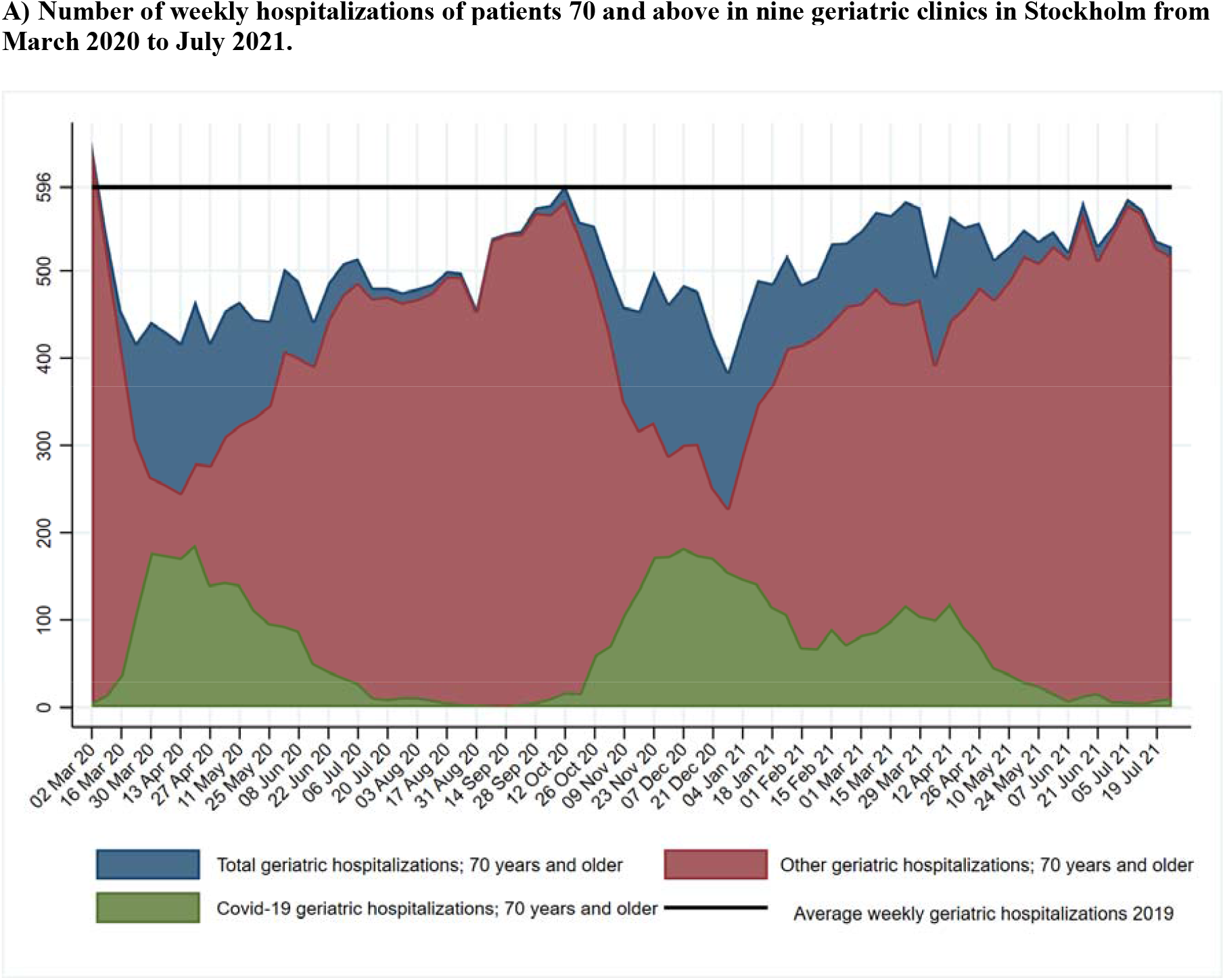
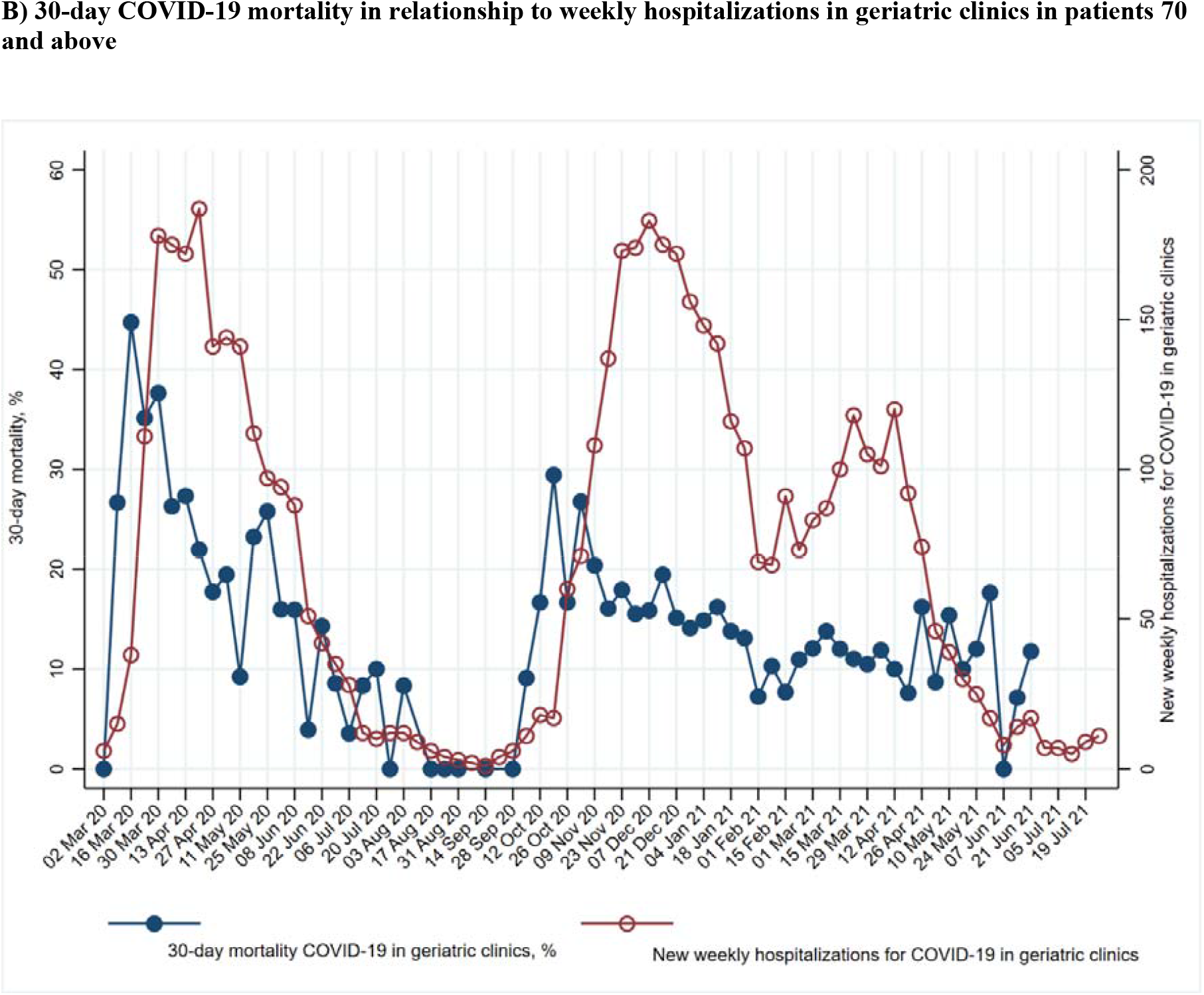
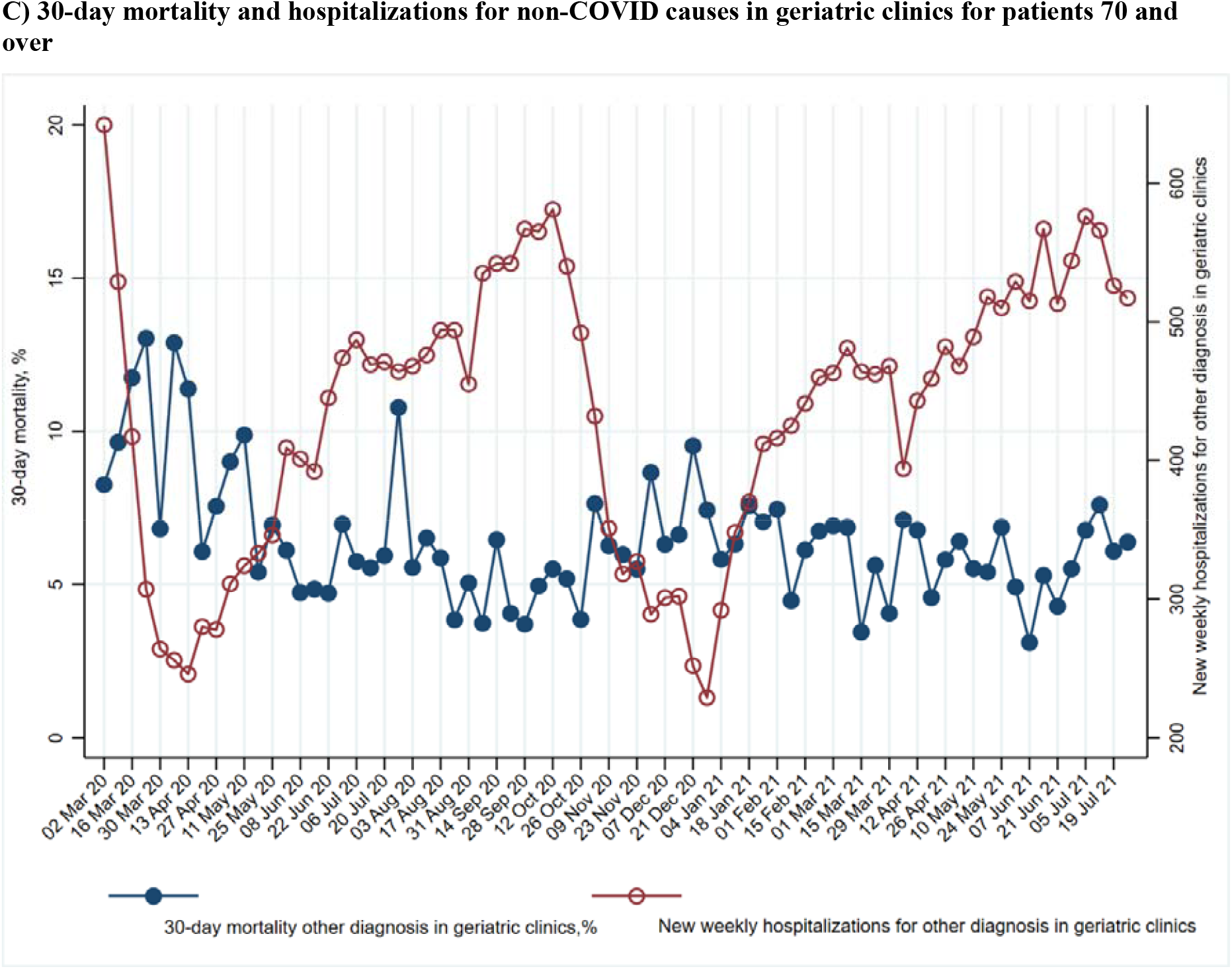
Hospitalizations and mortality throughout the COVID-19 pandemic for persons aged 70 and above.

### Thirty-day mortality

Thirty-day mortality was highest at the beginning of the first wave (29% in March-April 2020 for COVID-19; 10% for non-COVID-19), decreased as the first wave subsided (7% July-August for COVID-19; 6% for non-COVID-19), and increased again for COVID cases in the second wave (17% November-December for COVID; 7% for non-COVID). Thirty-day mortality remained more stable in the third wave (11 to 13% March-July 2021 for COVID-19; 6% non-COVID-19) (**Table 1**).

**Figure 2B** shows the relationship between weekly COVID-19 hospital admissions to the nine geriatric hospitals and 30-day mortality. Mortality is lowest when few COVID-19 patients are hospitalized. **Figure 2C** presents the same relationship but for other geriatric patients. The lowest mortality rates appear in the interpandemic peaks, when COVID hospitalizations were lowest and non-COVID hospitalizations were highest.

**Figure 3** shows the relationship between total COVID-19 cases and deaths in the Stockholm region and COVID-19 hospitalizations and deaths in the geriatric hospitals. The number of hospitalizations and the 30-day mortality rates increased with each pandemic peak and decreased between the peaks. The 30-day mortality rate after a positive test in Stockholm and the 30-day mortality rate after admission in the geriatric hospitals followed the pandemic and hospitalization curves. The smaller increase in the third wave probably indicates a vaccination effect.

**Figure 3.**
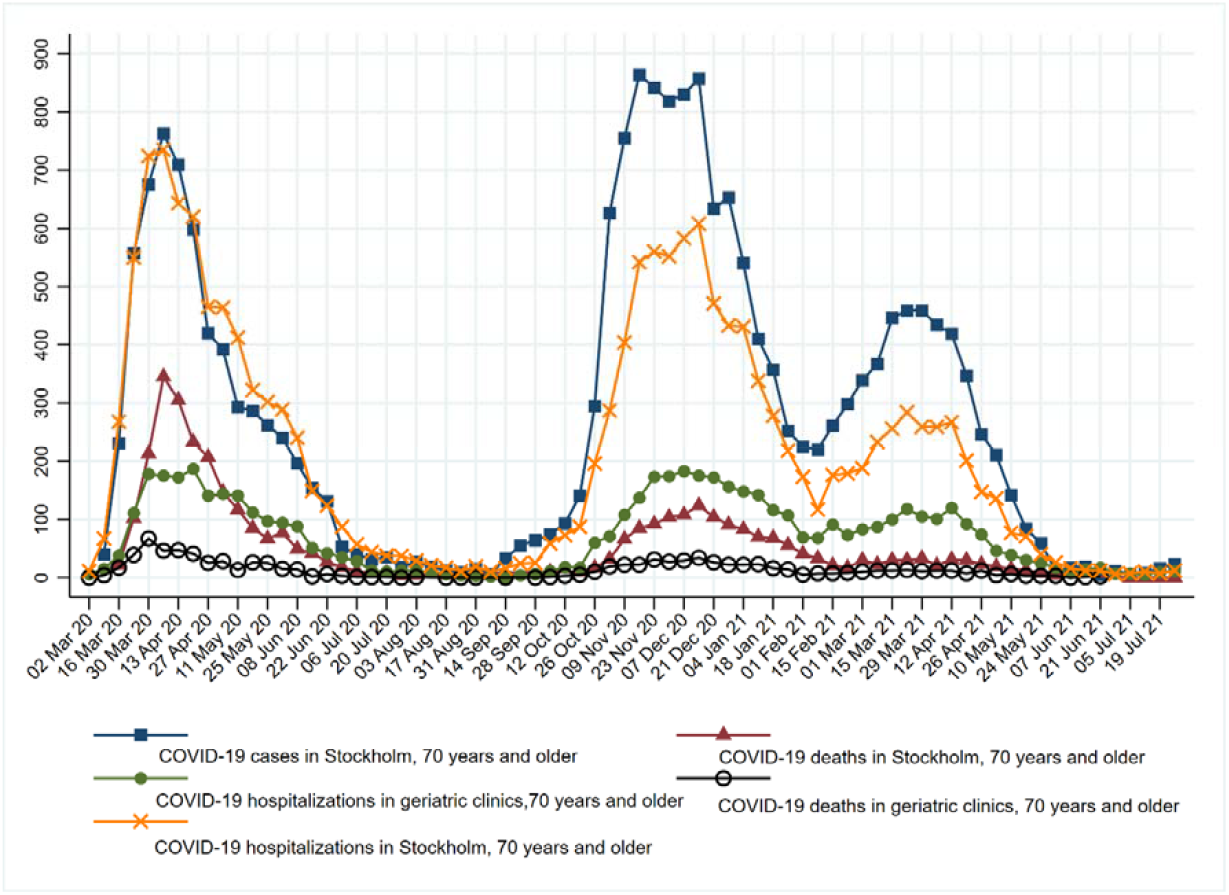
Relationship between confirmed cases of COVID-19, hospitalizations, and deaths in Stockholm.

**Table 2** shows 30-day mortality risk from logistic regression, adjusted by age, sex, Charlson Comorbidity Index (CCI) and treatment. Compared with patients admitted to geriatrics for COVID-19 in November-December 2020, the risk of 30-day mortality was higher (OR 1.92, 95% CI 1.55-2.37) in March-April 2020, and lower in July August 2020 (OR 0.31, 0.15-0.66). A downward tendency appeared again in January-February 2021(OR 0.70, 0.54-0.91), March-April 2021 (OR 0.72, 0.55-0.94) and May-June-July (OR 0.75, 0.49-1.13) (**Table 2**).

**Table 2.** Odds ratios for 30-day mortality in geriatric patients with COVID-19 and other diagnoses throughout the pandemic.

|  | COVID-19 |  | Non-COVID |  |
| --- | --- | --- | --- | --- |
|  | OR | 95%CI | OR | 95%CI |
| Mar-Apr 2020 | 1.92*** | 1.55,2.37 | 1.33** | 1.10,1.61 |
| May-June 2020 | 0.91 | 0.72,1.17 | 0.85 | 0.69,1.05 |
| July-Aug 2020 | 0.31** | 0.15,0.66 | 0.90 | 0.74,1.09 |
| Sep-Oct 2020 | 0.99 | 0.59,1.64 | 0.67*** | 0.55,0.82 |
| Nov-Dec 2020 | ref |  | ref |  |
| Jan-Feb 2021 | 0.70** | 0.54,0.91 | 0.93 | 0.76,1.14 |
| Mar-Apr 2021 | 0.72* | 0.55,0.94 | 0.81* | 0.67,0.99 |
| May-June-July 2021 | 0.75 | 0.49,1.13 | 0.82* | 0.69,0.98 |
| Age |  |  |  |  |
| 70-79 | ref |  | ref |  |
| 80-89 | 2.05*** | 1.68,2.51 | 1.62*** | 1.42,1.85 |
| 90+ | 3.06*** | 2.45,3.82 | 2.42*** | 2.11,2.77 |
| Women | 0.63*** | 0.54,0.74 | 0.68*** | 0.62,0.75 |
| CCI | 1.16*** | 1.12,1.20 | 1.18*** | 1.16,1.21 |
| ACEI | 0.94 | 0.78,1.14 | 0.84** | 0.75,0.94 |
| ARB | 0.84 | 0.70,1.01 | 0.72*** | 0.64,0.80 |
| β-blocker | 1.20* | 1.02,1.41 | 1.25*** | 1.13,1.39 |
| CCB | 0.80* | 0.68,0.95 | 0.72*** | 0.65,0.80 |
| Diuretics | 1.32*** | 1.12,1.55 | 1.49*** | 1.34,1.65 |
| Statins | 0.70*** | 0.59,0.82 | 0.74*** | 0.66,0.82 |
| Warfarin | 1.15 | 0.87,1.52 | 1.06 | 0.91,1.25 |
| LMWH | 1.06 | 0.88,1.26 | 1.23*** | 1.10,1.38 |
| NOAC | 0.93 | 0.77,1.13 | 0.98 | 0.88,1.08 |
| Glucocorticoids | 1.22* | 1.03,1.45 | 1.18** | 1.05,1.33 |
| Antibiotic | 1.51*** | 1.29,1.75 | 1.27*** | 1.16,1.40 |

Abbreviations: ACEI, angiotensin-converting enzyme inhibitors; ARB, angiotensin receptor blockers; CCB, calcium channel blockers; LMWH, low-molecular-weight heparin; NOAC, Non-vitamin-K antagonist oral anticoagulant.

### Thirty-day mortality in non-COVID-19 hospitalizations

Compared with hospitalizations in November-December 2020, hospitalizations in March-April 2020 had the highest mortality risk (OR 1.33; 95 CI 1.10-1.61), while hospitalizations from September-October 2020 (OR 0.67; 95% CI 0.55-0.82), March-April 2021 (0.81; 95% CI 0.67-0.99) and May-July 2021 (OR 0.82; 95% CI 0.69-0.98) presented lower risk, after adjusting (**Table 1, Figure 2**).

## DISCUSSION

Several important observations appear in this study of hospitalized geriatric patients in Stockholm, Sweden. First, hospital admissions fell sharply at the beginning of the pandemic and didn’t reach pre-pandemic levels throughout the whole observation period. Longer hospital stays in COVID-19 patients could be one reason for this finding. The length of hospitalization for COVID-19 patients was in line with previous international reports.^11, 14^ A reduction in elective procedures probably also contributed to the fall in admissions. The higher care burden for COVID-19 patients, together with the time required to don and doff protective equipment may have led to greater care burden and lower capacity to accept new cases. Additionally, hip and other osteoporotic fractures might have decreased in the population 70 and older due to the recommendations for this age group to avoid social contact.^15^ Hospital admissions further decreased during the summer months: the receding COVID-19 waves probably contributed to this reduction as did the customary reductions in available beds due to staff vacation which occur every summer.

Care seeking behaviour and the capacity of the healthcare system to detect and manage other diseases was affected during the pandemic: for example, the Swedish Stroke Register reports a decrease in the number of detected strokes in 2020 and 2021 compared to 2019.^16^. The Swedish registry for cognitive/dementia disorders reported a 30% reduction in diagnostic work-up.^17^ One US study included over 23 million emergency department (ED) visits for elderly Medicare beneficiaries, and compared hospital admission rate and changes in 30-day mortality between identical periods in 2018-19 and 2019-20.^18^ The drop in ED visits and increase in admission rate didn’t correlate with local COVID-19 incidence. Mortality risk after ED visits also increased. The drop in ED visits, together with increased admission rate and mortality suggest that patients were avoiding seeking care, selecting for a sicker patient population in the ED. Furthermore, the lack of correlation with COVID-19 incidence suggests that patient behaviours (rather than hospital capacity) were responsible.^18^ Another US study found a decrease in ED visits for serious cardiovascular conditions early in the pandemic with gradual recovery (until October 2020)^19^. The same drop in cardiovascular visits was seen in Europe.^20, 21^ In contrast to the US study we saw a drop in hospitalizations in our cohort but not a complete recovery afterwards. We do not have data on ED visits so we cannot distinguish between patient care-seeking behaviours and triage in the ED. As seen in figure 2A COVID-19 patients displaced other diagnoses from geriatric care. The supply of geriatric hospitalizations in our cohort was inelastic, remained relatively stable during the pandemic and was lower than in 2019.

Second, 30-day mortality rose and fell with the pandemic waves. However, the peak mortality in geriatric clinics was higher in the first than in the second wave. This effect was more evident for COVID-19 patients but appeared also in non-COVID geriatric patients. Several studies, including our previous study, have noted a decrease in mortality throughout the first wave of the pandemic^22 1^. A subsequent increase was observed again during the second wave in Sweden ^9, 23^. One study included more than 30,000 COVID-19 patients (including those admitted to the ICU) hospitalized between March and December 2020 in Sweden and showed that the 60-day case fatality rate for hospitalized cases decreased in the first wave but increased again in the second wave^9^. This study noted that the average age and comorbidities for hospitalized patients increased in the second wave, although they adjusted for these and other factors.

COVID-19 incidence was more severely underreported in the first wave in Sweden than later in the pandemic.^23^ Some patients were treated in their nursing homes and never transferred to hospital, and this could have the effect of lowering the average age and comorbidities in the first wave but, interestingly, this change in average age is not present in our cohort. This may be explained by the fact that most patients admitted to inpatient geriatric hospital care lived in their own homes prior to admission. In this geriatric cohort, the proportion of patients presenting with oxygen saturation under 90% at baseline decreased from 8.9% in March-April 2020 to 2% in June, increasing again to 7.7% in December 2020. In our previous article we showed that low oxygen saturation at baseline explained some, but not all, of the temporal trend in mortality observed during the first wave.^1^ Triage was probably harsher in the first wave leading to a sicker patient population and higher mortality. The alpha variant arrived in Sweden in December 2020, too late to explain the increase in 30-day mortality. Improvements in treatment may also explain a part of the reduction in mortality.

In an international context, studies from Africa^24^, Italy^25^, and Germany^26^ showed a significant difference in COVID-mortality between the first wave and the second wave. Our temporal trend closely follows the German report, with high initial case fatal ratio, nadir in summer 2020 and subsequent increase in the second wave. In Germany, the first wave was smaller and led to fewer hospitalizations than the second, while the peaks in hospitalizations between the waves were more similar in Sweden. Despite this, the highest mortality in both studies was seen in the initial peak of the first wave^26^. Another study examined raw case fatality ratio (CFR) in 53 countries or regions and showed decreased mortality in the second wave in 43 of them, including in Sweden.^27^

Third, vaccinations probably changed the association between incidence and mortality in the third wave and averted the increase in 30-day mortality that had been apparent with increasing hospitalizations in the previous waves. The COVID-19 vaccination campaign began at the end of December 2020 in nursing homes and its effects on mortality were felt already in March-April 2021, as also seen in our cohort.^28^

We cannot completely explain the changes in mortality. First, it should be noted that the 30-day mortality rate for COVID-19 patients reported in our study is based on a large cohort of older people in geriatric clinics in Stockholm, excluding nursing home patients treated in place, mild cases treated at home or cases treated in ICU or infectious disease wards instead of the geriatric clinics. Second, during the first wave of pandemic, the treatment and care of COVID-19 patients underwent great changes (https://www.internetmedicin.se/behandlingsoversikter/infektion/covid-19/), and these improvements are likely to be part of the reason for the decline in mortality during the first wave. In the second wave, the national guidelines for COVID-19 patients hospitalized in Sweden didn’t undergo major changes. In a previous report, we have shown that the prognosis of COVID-19 for the most frail during the first wave was ominous ^29^. It is possible that the most vulnerable and frail died during the first wave and thus contributing to lower mortality during the second wave.

We believe that due to the inelasticity of hospitalizations, increasing COVID-19 admissions during pandemic waves led to corresponding decreases in non-COVID admissions. This may have led to more stringent selection of admissions, a sicker patient population and greater overall mortality. Changes in treatment guidelines, particularly use of anticoagulants and corticosteroids led to improvement in the outcome of patients with COVID-19^30^. However, except for vaccines and the third wave, these changes were implemented relatively early in the pandemic (before or during the summer of 2020) and cannot explain all the later changes in mortality.^30^

### Strengths and limitations

In Sweden, geriatric hospitalizations are indicated on criteria of biological (and not chronological aging). Non-frail and non-comorbid older patients may have been hospitalized in other clinics (eg. infectious diseases) and not included in our cohort. Nine out of eleven existing geriatric clinics in Stockholm participated in the study. Living situation, comorbidities and medications were obtained from electronic health records with imperfect ascertainment. Medications were considered present if they were present or prescribed within 24h of hospitalization and would have included both newly prescribed medications and those removed after admission. We did not have information on vaccination, which would have been useful for assessing the decline of mortality in the third wave of the pandemic. Strengths of this study are the large geriatric cohort including nine out of eleven geriatric clinics in Stockholm treating patients with COVID-19. Also novel is the analysis of geriatric hospitalizations for other causes and the long study period including three pandemic waves over a period of 17 months.

## Conclusion

Thirty-day mortality was highest at the peak of the first wave, decreased in the inter-wave period and then increased again but to a lower peak in the second wave. The mortality increase of the third wave was probably averted by the vaccination campaign. Non-COVID mortality showed a similar trend but with lower magnitude. During COVID-19 incidence peaks, COVID-19 hospitalizations displaced non-COVID geriatric patients. Despite a massive organizational effort and the individual effort and sacrifice of health care workers, the healthcare system could not compensate for high community spread of COVID-19 during the pandemic peaks. Hospital admissions fell at the beginning of the pandemic and never returned to pre-pandemic levels, probably reflecting the greater complexity of COVID-19 patients compared to ordinary geriatric patients. The sustained fall in hospital admissions is particularly worrisome considering the backlog of cancelled and deferred care caused by the pandemic. COVID-19 disproportionally affects geriatric patients, and the broader pandemic has disrupted geriatric care.

## Data Availability

All data produced in the present study are available upon reasonable request to the authors subject to the relevant provisions for personal data management of Swedish and European legislation

## ACKNOWLEDGEMENTS

Sara Garcia-Ptacek has received funding from the Swedish Stroke Association, the regional agreement on medical training and clinical research between the Stockholm County council and the Karolinska Institutet (ALF) and FORTE (**#**2017-01646). Hong Xu by StratNeuro (the Strategic Research Area Neuroscience-Karolinska Institutet, Umeå University and KTH) and the Center for Innovative Medicine (CIMED). Maria Eriksdotter by the Swedish medical research council grant (#2016-02317 and #2020-02014) and the regional agreement on medical training and clinical research between the Stockholm County council and the Karolinska Institutet (ALF), Dorota Religa by the Swedish Research Council (#2012-2291 and 2020-06101). The authors are grateful to the Swedish Board of Health and Welfare (Frida Broström and Henrik Nordin) for providing epidemiological data on COVID-19 in the population 70 years and older in Stockholm. The authors appreciate covidxix.org for their clear presentation of Swedish Covid-19 statistics and their help in finding relevant data for this study.

---

**eTable 1.** COVID-19 cases, mortality and hospitalizations in Stockholm and in geriatric clinics for population 70 and above.

| Week | COVID-19 positive cases in Stockholm | 30-day COVID-19 mortality Stockholm | COVID-19 hospitalizations in Stockholm | Total hospitalizations in Stockholm | COVID-19 geriatric hospitalizations | 30-day mortality after geriatric hospitalization for COVID-19 | Other diagnoses, geriatric hospitalizations | 30-day mortality after geriatric hospitalization, non-COVID-19 | Total geriatric hospitalizations |
| --- | --- | --- | --- | --- | --- | --- | --- | --- | --- |
| March 1, 2020 |  |  |  |  |  |  | 28 | 3 | 28 |
| March 2, 2020 |  |  | 11 | 2 718 | 6 | 0 | 642 | 53 | 648 |
| March 9, 2020 | 40 | X | 67 | 2 515 | 15 | 4 | 529 | 51 | 544 |
| March 16, 2020 | 231 | 22 | 268 | 2 364 | 38 | 17 | 417 | 49 | 455 |
| March 23, 2020 | 557 | 102 | 550 | 2 237 | 111 | 39 | 307 | 40 | 418 |
| March 30, 2020 | 675 | 213 | 724 | 2 130 | 178 | 67 | 264 | 18 | 442 |
| April 6, 2020 | 763 | 345 | 735 | 2 120 | 175 | 46 | 256 | 33 | 431 |
| April 13, 2020 | 709 | 305 | 643 | 2 116 | 172 | 47 | 246 | 28 | 418 |
| April 20, 2020 | 598 | 233 | 620 | 2 260 | 187 | 41 | 280 | 17 | 467 |
| April 27, 2020 | 420 | 207 | 465 | 2 030 | 141 | 25 | 278 | 21 | 419 |
| May 4, 2020 | 392 | 147 | 464 | 2 245 | 144 | 28 | 311 | 28 | 455 |
| May 11, 2020 | 293 | 117 | 412 | 2 120 | 141 | 13 | 324 | 32 | 465 |
| May 18, 2020 | 286 | 85 | 322 | 1 999 | 112 | 26 | 333 | 18 | 445 |
| May 25, 2020 | 262 | 67 | 302 | 2 186 | 97 | 25 | 346 | 24 | 443 |
| June 1, 2020 | 240 | 77 | 289 | 2 277 | 94 | 15 | 409 | 25 | 503 |
| June 8, 2020 | 196 | 49 | 240 | 2 320 | 88 | 14 | 401 | 19 | 489 |
| June 15, 2020 | 154 | 42 | 149 | 2 081 | 51 | 2 | 392 | 19 | 443 |
| June 22, 2020 | 131 | 27 | 124 | 2 235 | 42 | 6 | 445 | 21 | 487 |
| June 29, 2020 | 53 | 18 | 87 | 2 187 | 35 | 3 | 474 | 33 | 509 |
| July 6, 2020 | 40 | 11 | 57 | 2 125 | 28 | 1 | 487 | 28 | 515 |
| July 13, 2020 | 31 | 7 | 44 | 2 018 | 12 | 1 | 469 | 26 | 481 |
| July 20, 2020 | 35 | 7 | 38 | 2 010 | 10 | 1 | 471 | 28 | 481 |
| July 27, 2020 | 18 | 4 | 37 | 2 078 | 12 | 0 | 464 | 50 | 476 |
| August 3, 2020 | 27 | 5 | 29 | 2 136 | 12 | 1 | 468 | 26 | 480 |
| August 10, 2020 | 19 | X | 21 | 2 145 | 9 | x | 476 | 31 | 485 |
| August 17, 2020 | 12 | 5 | 17 | 2 336 | 6 | 0 | 494 | 29 | 500 |
| August 24, 2020 | 9 | X | 11 | 2 453 | 4 | 0 | 494 | 19 | 498 |
| August 31, 2020 | 15 | X | 19 | 2 420 | 3 | 0 | 455 | 23 | 458 |
| September 7, 2020 | 6 | X | 10 | 2 552 | 2 | x | 535 | 20 | 537 |
| September 14, 2020 | 33 | X | 16 | 2 567 | 1 | 0 | 542 | 35 | 543 |
| September 21, 2020 | 55 | X | 24 | 2 552 | 4 | x | 542 | 22 | 546 |
| September 28, 2020 | 64 | X | 25 | 2 673 | 6 | 0 | 567 | 21 | 573 |
| October 5, 2020 | 74 | 10 | 59 | 2 726 | 11 | 1 | 565 | 28 | 576 |
| October 12, 2020 | 93 | X | 73 | 2 655 | 18 | 3 | 581 | 32 | 599 |
| October 19, 2020 | 141 | 10 | 87 | 2 592 | 17 | 5 | 540 | 28 | 557 |
| October 26, 2020 | 295 | 17 | 196 | 2 683 | 60 | 10 | 492 | 19 | 552 |
| November 2, 2020 | 626 | 32 | 287 | 2 545 | 71 | 19 | 432 | 33 | 503 |
| November 9, 2020 | 755 | 66 | 404 | 2 433 | 108 | 22 | 351 | 22 | 459 |
| November 16, 2020 | 864 | 85 | 542 | 2 565 | 137 | 22 | 318 | 19 | 455 |
| November 23, 2020 | 842 | 92 | 560 | 2 579 | 173 | 31 | 327 | 18 | 500 |
| November 30, 2020 | 818 | 104 | 552 | 2 596 | 174 | 27 | 289 | 25 | 463 |
| December 7, 2020 | 830 | 108 | 583 | 2 680 | 183 | 29 | 301 | 19 | 484 |
| December 14, 2020 | 857 | 124 | 608 | 2 590 | 175 | 34 | 302 | 20 | 477 |
| December 21, 2020 | 634 | 104 | 471 | 2 247 | 172 | 26 | 252 | 24 | 424 |
| December 28, 2020 | 653 | 91 | 433 | 2 308 | 156 | 22 | 229 | 17 | 385 |
| January 4, 2021 | 540 | 83 | 431 | 2 361 | 148 | 22 | 292 | 17 | 440 |
| January 11, 2021 | 410 | 70 | 338 | 2 501 | 142 | 23 | 348 | 22 | 490 |
| January 18, 2021 | 357 | 68 | 278 | 2 512 | 116 | 16 | 370 | 28 | 486 |
| January 25, 2021 | 252 | 56 | 218 | 2 475 | 107 | 14 | 412 | 29 | 519 |
| February 1, 2021 | 225 | 41 | 173 | 2 403 | 69 | 5 | 416 | 31 | 485 |
| February 8, 2021 | 220 | 33 | 117 | 2 363 | 68 | 7 | 425 | 19 | 493 |
| February 15, 2021 | 261 | 21 | 176 | 2 513 | 91 | 7 | 441 | 27 | 532 |
| February 22, 2021 | 298 | 17 | 179 | 2 504 | 73 | 8 | 460 | 31 | 533 |
| March 1, 2021 | 339 | 30 | 188 | 2 446 | 83 | 10 | 463 | 32 | 546 |
| March 8, 2021 | 367 | 23 | 233 | 2 576 | 87 | 12 | 481 | 33 | 568 |
| March 15, 2021 | 446 | 30 | 256 | 2 510 | 100 | 12 | 464 | 16 | 564 |
| March 22, 2021 | 459 | 30 | 284 | 2 664 | 118 | 13 | 462 | 26 | 580 |
| March 29, 2021 | 459 | 33 | 259 | 2 337 | 105 | 11 | 468 | 19 | 573 |
| April 5, 2021 | 434 | 21 | 259 | 2 357 | 101 | 12 | 394 | 28 | 495 |
| April 12, 2021 | 419 | 31 | 267 | 2 435 | 120 | 12 | 443 | 30 | 563 |
| April 19, 2021 | 347 | 30 | 201 | 2 376 | 92 | 7 | 459 | 21 | 551 |
| April 26, 2021 | 246 | 23 | 147 | 2 463 | 74 | 12 | 482 | 28 | 556 |
| May 3, 2021 | 211 | 19 | 136 | 2 529 | 46 | 4 | 468 | 30 | 514 |
| May 10, 2021 | 142 | 15 | 77 | 2 318 | 39 | 6 | 489 | 27 | 528 |
| May 17, 2021 | 84 | 13 | 71 | 2 582 | 30 | 3 | 518 | 28 | 548 |
| May 24, 2021 | 59 | 8 | 40 | 2 469 | 25 | 3 | 510 | 35 | 535 |
| May 31, 2021 | 12 | 5 | 26 | 2 537 | 17 | 3 | 529 | 26 | 546 |
| June 7, 2021 | 17 | X | 16 | 2 462 | 8 | 0 | 515 | 16 | 523 |
| June 14, 2021 | 19 | X | 12 | 2 553 | 14 | 1 | 567 | 30 | 581 |
| June 21, 2021 | 5 | X | 12 | 2 289 | 17 | 2 | 513 | 22 | 530 |
| June 28, 2021 | 11 | X | 6 | 2 362 | 7 | x | 544 | 30 | 551 |
| July 5, 2021 | 6 | 0 | 7 | 2 271 | 7 | x | 576 | 39 | 583 |
| July 12, 2021 | 8 | 0 | 10 | 2 107 | 5 | x | 566 | 43 | 571 |
| July 19, 2021 | 16 | 0 | 7 | 2 105 | 9 | x | 526 | 32 | 535 |
| July 26, 2021 | 23 | 0 | 12 | 2 134 | 11 | x | 517 | 33 | 528 |

